# Smoking Habits and the Onset of Stroke: Insights from the Kyoto Stroke Registry

**DOI:** 10.1101/2023.09.07.23295229

**Authors:** Kazuo Shigematsu, Hiromi Nakano, the Kyoto Stroke Registry Committee

## Abstract

Cigarette smoking is a well-established risk factor for stroke due to its contribution to atherosclerosis. This study utilized data from the Kyoto Stroke Registry (KSR) spanning April 1997 to March 2020 to investigate the relationship between smoking history and the age at which stroke occurs.

The study focused on stroke patients registered in the KSR, categorizing them into four groups based on their smoking history: those with no smoking history, individuals who quit smoking more than one year ago, those who smoked fewer than 20 cigarettes per day, and those who smoked more than 20 cigarettes per day. The primary objective was to compare the age at which stroke onset occurred among these groups, employing statistical analysis through a Student t-test to identify significant differences.

After excluding patients with missing smoking status information, the analysis involved 30,491 stroke patients. The results revealed a noteworthy association: individuals with a smoking history, particularly heavy smokers, experienced a significantly younger age at stroke onset compared to those who had never smoked. Moreover, the study found that quitting smoking for more than one year was linked to a later onset of stroke compared to continuing to smoke. This observation held true even among those who smoked less than 20 cigarettes per day. Furthermore, the study’s findings remained consistent across all stroke types, including cerebral infarction, cerebral hemorrhage, and subarachnoid hemorrhage, regardless of gender.

In conclusion, this comprehensive analysis of the KSR data reinforces the well-established notion that smoking history plays a critical role in the timing of stroke onset. Importantly, it underscores the potential benefits of smoking cessation, as well as the avoidance of smoking altogether, in reducing the risk of stroke. These findings carry significant implications for public health initiatives aimed at stroke prevention and emphasize the pivotal role of smoking cessation programs in mitigating the risk of stroke-associated morbidity and mortality.

## Introduction

Cigarette smoking is a well-known cause of atherosclerosis and a recognized risk factor for stroke ^1^. The Kyoto Stroke Registry (KSR) is an ongoing study commissioned by the Kyoto Medical Association, focusing on stroke incidence in Kyoto Prefecture. The registry includes detailed information on patients’ smoking history. Participants were categorized into four groups: no smoking history, quit smoking more than one year ago, smoked less than 20 cigarettes per day, and smoked more than 20 cigarettes per day. The aim was to compare the age of stroke onset among these groups. We hypothesized that if the smoking group is more prone to stroke than the non-smoking group, then the stroke onset would occur at a younger age.

## Method

We investigated the relationship between smoking history and age at stroke onset in patients registered with the KSR between April 1997 and March 2020. Detailed information on the KSR has been previously reported^2-6^. In summary, the KSR collaborates with all medical institutions affiliated with the Kyoto Medical Association to comprehensively study stroke patients in Kyoto Prefecture. The data are analyzed to improve stroke prevention, rehabilitation, and overall patient outcomes with the goal of minimizing hospitalization and facilitating return to normal life. The registration form is distributed to all cooperating facilities by the Kyoto Medical Association, and the responsible physicians at these facilities complete the form and return it to the association. The data are compiled by the Medical Association. Stroke cases were classified into three major subtypes: cerebral infarction (CI), cerebral hemorrhage (CH), and subarachnoid hemorrhage (SAH), following the WHO definition ^7^. This research was conducted in accordance with the ethical principles outlined in the Declaration of Helsinki for medical research involving human subjects. Approval was obtained from the Board of Directors of the Kyoto Medical Association, the Department of Health and Welfare of Kyoto Prefecture, and the Ethics Committee of the Japan Medical Association. Since all personally identifiable information was removed from the secondary files before analysis, the requirement for written informed consent from the patients was waived by the respective boards.

During the study period, a total of 36,303 patients were registered, and after excluding 5,812 patients with missing smoking status information, 30,491 patients were included in the analysis. Table 1 provides a summary of the study population.

**Table 1:** Patient registration by smoking history and stroke subtype.

|  | CI | CH | SAH | CI+CH | CI +<br>SAH | CH +<br>SAH | Male<br>and<br>female<br>(% male<br>female) |
| --- | --- | --- | --- | --- | --- | --- | --- |
| No<br>history of<br>smoking | 14090 | 4473 | 1278 | 29 | 7 | 8 | 7705 ·<br>12180<br>(38.7 ·<br>61.3) |
| Quit<br>smoking<br>more<br>than 1<br>year ago | 2442 | 573 | 77 | 8 | 0 | 3 | 2729 ·<br>374<br>(87.9 ·<br>12.1) |
| Smoked<br>less than<br>20<br>cigarettes<br>per day | 3708 | 992 | 462 | 8 | 3 | 8 | 4152 ·<br>1029<br>(80.1 ·<br>19.9%) |
| Smoked<br>more<br>than 20<br>cigarettes<br>per day | 1640 | 488 | 181 | 6 | 2 | 5 | 2027 ·<br>243<br>(89.5 ·<br>10.5) |

CI: cerebral infarction. CH: cerebral hemorrhage. SAH: subarachnoid hemorrhage.

Patients were divided into four counties according to their cigarette smoking history (1) no smoking history 2) quit smoking for at least one year 3) smoking less than 20 cigarettes per day 4) smoking more than 20 cigarettes per day). Smoking history information was obtained from the patient, family members, and/or medical records.

The age of stroke onset for each group was compared. A Student t-test was used to determine if there was a difference in age of onset between groups. Furthermore, we examined whether there was a difference in age at onset for the three main stroke types: cerebral infarction, cerebral hemorrhage, and subarachnoid hemorrhage, depending on the smoking history of the patients. Finally, to determine whether these differences differed by gender, we examined the age of onset for each of the three types of stroke, by gender, and by the four smoking history groups. All personal information was removed from the analysis, and no personal information was included in the results. Since the purpose of the registry is to promote public health, written consent was not obtained from individuals, and the continuation of the registry without such consent is permitted by the government. This study was reviewed and approved by the Ethics Committee of the Japan Medical Association.

Patients were categorized into four groups based on their cigarette smoking history: 1) no smoking history, 2) quit smoking for at least one year, 3) smoking less than 20 cigarettes per day, and 4) smoking more than 20 cigarettes per day. Smoking history information was obtained from the patients themselves, their family members, and/or medical records. The age of stroke onset was compared among these groups. To assess differences in age of onset between groups, a Student t-test was employed. Additionally, we examined whether there were variations in age at onset for the three primary stroke types (cerebral infarction, cerebral hemorrhage, and subarachnoid hemorrhage) based on the smoking history of the patients. Furthermore, we explored gender differences by examining the age of onset for each stroke type, stratified by gender and smoking history groups. Personal information was completely anonymized during the analysis, and the results did not contain any identifying information. Given that the registry aims to promote public health, written consent was not obtained from individuals, and the continuation of the registry without such consent is permitted by the government. This study received approval from the Ethics Committee of the Japan Medical Association.

## Results

To determine if there were significant differences in the age at stroke onset among the groups, we examined the mean, standard deviation, and mean age at stroke onset for each of the four defined groups based on smoking habits and history. Cases with multiple stroke types, such as CI + CH, were excluded from the statistical analysis when comparing by stroke type. The results are presented in Table 2. The age of onset for the four groups was as follows: 75.20, 73.74, 66.58, and 62.46 years, respectively. The age of onset decreased significantly with higher levels of smoking. Smoking cessation for more than one year was associated with a significantly later onset of stroke compared to continued smoking, even for individuals smoking less than 20 cigarettes per day, although their onset was still younger than that of non-smokers.

**Table 2.**
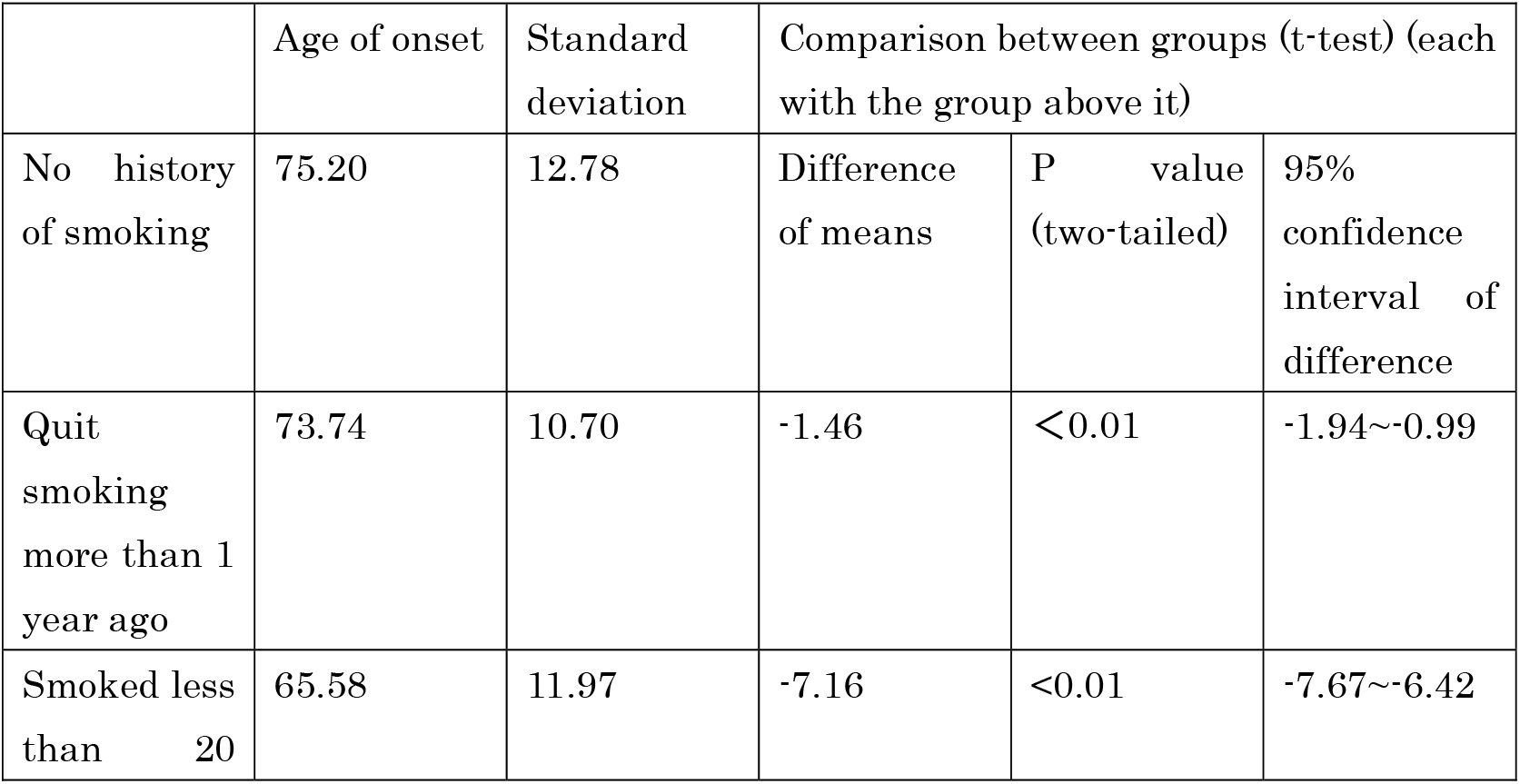
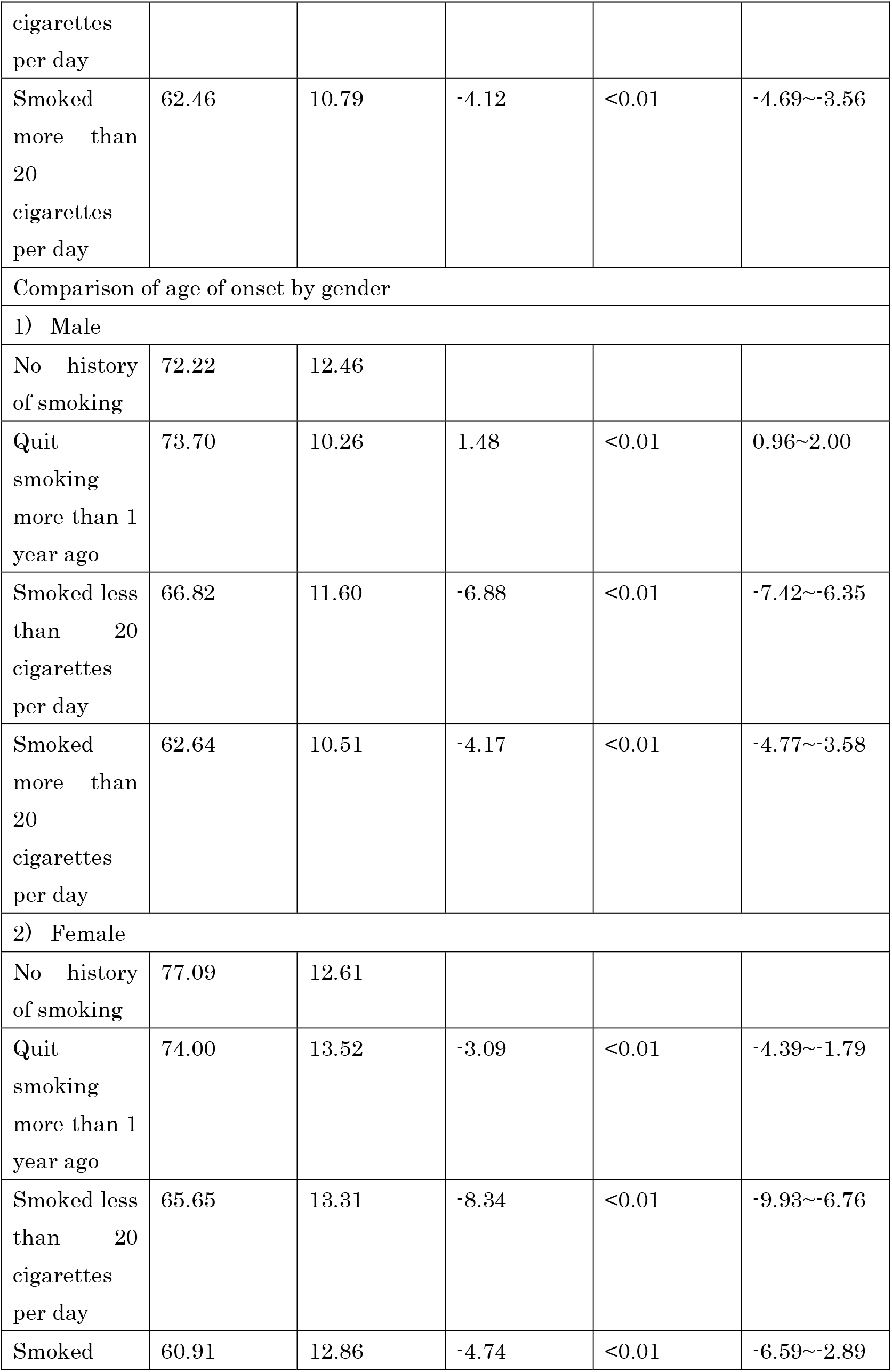
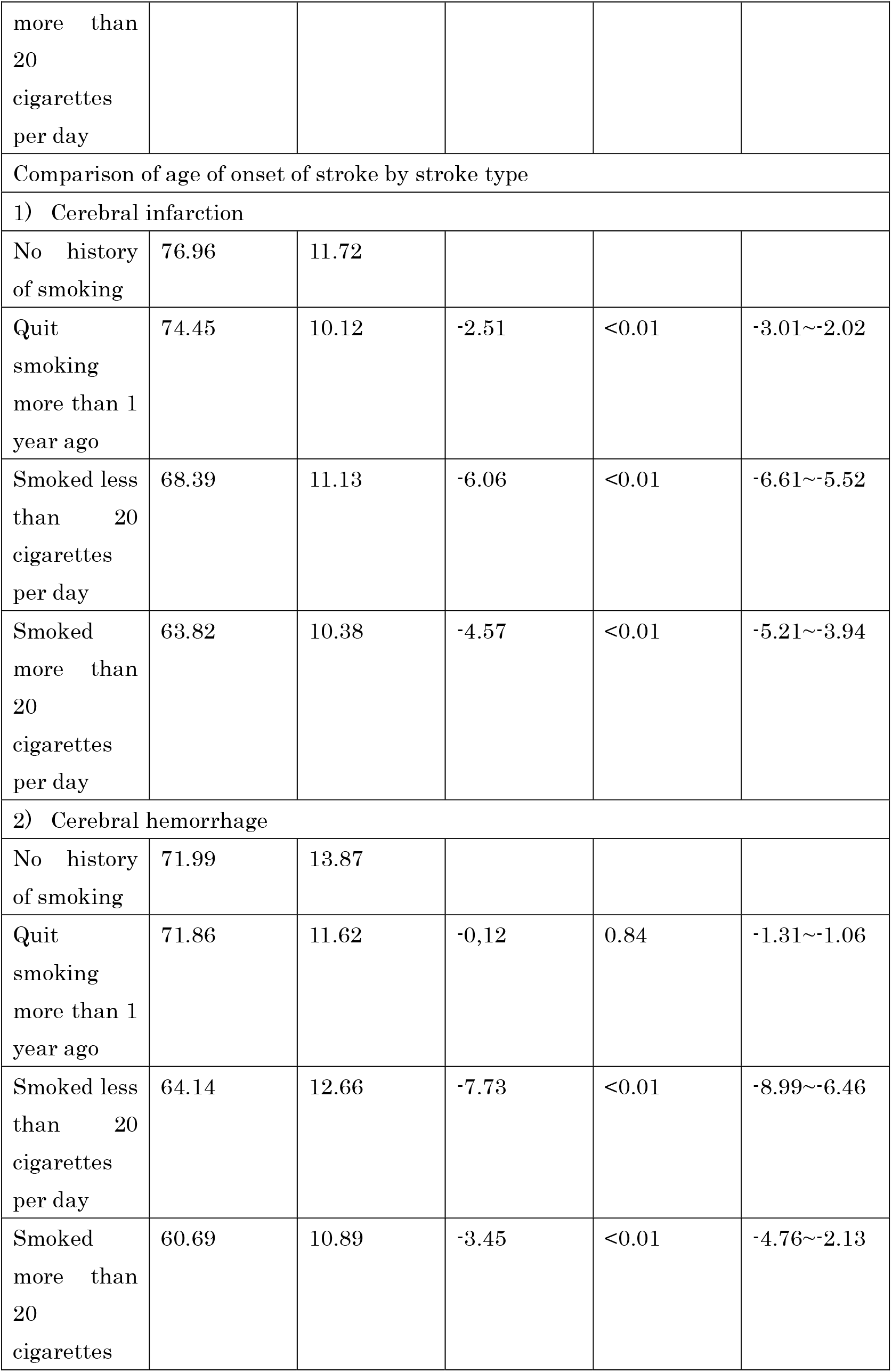
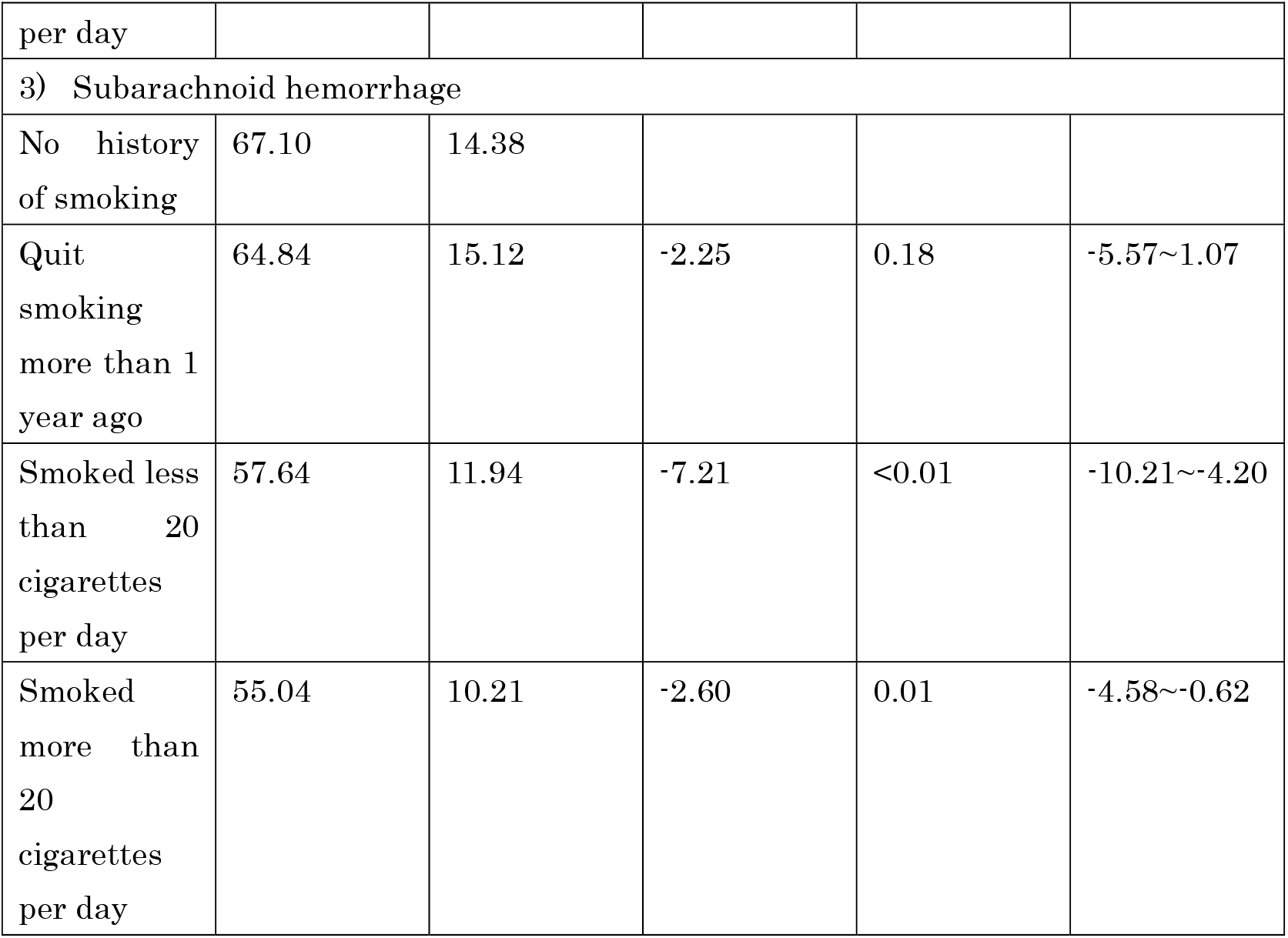
Comparison of stroke onset ages by smoking history.

Furthermore, we conducted comparisons within each stroke type: CI, CH, and SAH. The age of onset for CI, in order of the groups, was 79.69, 74.45, 68.39, and 63.82 years. For CH, the respective ages of onset were 71.99, 71.86, 64.14, and 60.69 years. Lastly, for SAH, the ages of onset were 67.10, 64.84, 57.64, and 55.04 years.

Considering the considerable differences in smoking habits between men and women in Japan, we conducted separate analyses for each gender. For the overall incidence of stroke, the order of age at onset among men was as follows: 72.22, 73.70, 66.82, and 62.64 years. Similarly, for women, the ages of onset were 77.09, 74.00, 65.65, and 60.91 years.

Table 2 provides a summary of the results obtained from the analysis.

## Discussion

In this study, we have epidemiologically demonstrated that smoking accelerates the age of onset of stroke.

Individuals with a history of smoking exhibited a younger age of stroke onset compared to those without such a history. This pattern held true for both genders and across all stroke types: cerebral infarction, cerebral hemorrhage, and subarachnoid hemorrhage. This implies that smoking is a risk factor for all stroke types and genders. For instance, those who smoked 20 or more cigarettes per day experienced stroke onset roughly 13 years earlier than non-smokers. In essence, if they had abstained from smoking, the onset of stroke could have been delayed by nearly 13 years. Stroke not only poses a risk to life but is also associated with potential complications like paralysis, aphasia, and cognitive impairment. A stroke occurring in one’s mid-60s, when they might otherwise be in good health and able to work or enjoy life, can drastically impact their quality of life and work capacity.

The age of stroke onset for individuals who quit smoking for at least a year was older than for those who continued smoking, suggesting that smoking cessation may delay the onset of stroke.

Smoking is linked to an increased risk of stroke, and the mechanisms behind this association include:

a. Atherosclerosis^8^: Smoking significantly contributes to the development and progression of atherosclerosis, a key mechanism in ischemic stroke pathogenesis. Tobacco smoke constituents, including nicotine and carbon monoxide, lead to endothelial dysfunction, induce inflammation, and contribute to the formation of atherosclerotic plaques within cerebral vessels. Consequently, the risk of ischemic stroke is elevated.
b. Thrombosis and platelet aggregation^9^: Smoking enhances platelet activation and aggregation, creating a pro-thrombotic environment. This heightened platelet reactivity is a significant risk factor for ischemic stroke as it can lead to thrombus formation within cerebral blood vessels, potentially causing occlusion.
c. Hypertension: Smoking is a major contributor to hypertension, a well-established risk factor for stroke. Nicotine, a prominent component of tobacco products, stimulates the release of adrenaline and narrows blood vessels, leading to elevated blood pressure^10^. This sustained increase in blood pressure further amplifies the risk of both ischemic and hemorrhagic strokes.
d. Oxidative stress^11^: Smoking induces oxidative stress, characterized by an imbalance between reactive oxygen species (ROS) and antioxidants. This oxidative stress negatively affects the vascular endothelium, causing damage and dysfunction. Consequently, cerebral vessels become more vulnerable to atherosclerosis and thrombosis, increasing the risk of stroke.
e. Hemorrhagic stroke^12, 13^: It is noteworthy that smoking also increases the risk of hemorrhagic stroke, although the precise mechanisms are still under investigation. One possible explanation lies in the adverse effects of smoking on blood vessel integrity. Smoking may weaken vessel walls and contribute to increased blood pressure, rendering cerebral vessels more susceptible to rupture.

Understanding these intricate mechanisms not only highlights the detrimental impact of smoking on stroke risk but also underscores the importance of promoting smoking cessation as a critical preventive measure to alleviate the burden of stroke on public health. Further research into the precise molecular pathways involved in these mechanisms may provide insights into targeted interventions for stroke prevention among smokers.

### Limitations

Collecting accurate and quantitative information on lifestyle habits, such as the number of cigarettes smoked per day, can be challenging. Additionally, various factors such as gender differences in smoking rates, health consciousness, doctor guidance, working hours, and income might confound the analysis. Despite these limitations, due to the extensive dataset and the volume-dependent impact of smoking on stroke, the study’s conclusion that smoking accelerates stroke onset is likely robust.

## Conclusions

Data from the Kyoto Stroke Registry revealed that patients with a smoking history, especially heavier smokers, experienced a younger age of stroke onset compared to non-smokers. Smoking cessation led to a stroke onset age closer to that of non-smokers than to that of continuing smokers. These trends persisted regardless of gender and were consistent across all stroke types: cerebral hemorrhage and subarachnoid hemorrhage. These findings indicate that avoiding smoking or quitting smoking can be effective in preventing strokes.

## Data Availability

Data is managed by the Kyoto Medical Association. Please submit a request for access, including your reasons. If your request is approved, you will be granted access.

## References

1. Shinton R, Beevers G. Meta-analysis of relation between cigarette smoking and stroke. British Medical Journal. 1989;298:789–794

2. Shigematsu K, Watanabe Y, Nakano H, Committee KSR. Influences of hyperlipidemia history on stroke outcome; a retrospective cohort study based on the kyoto stroke registry. BMC neurology. 2015;15:1–6

3. Shigematsu K, Watanabe Y, Nakano H, Committee KSR. Weekly variations of stroke occurrence: An observational cohort study based on the kyoto stroke registry, japan. BMJ open. 2015;5:e006294

4. Shigematsu K, Nakano H, Watanabe Y, Sekimoto T, Shimizu K, Nishizawa A, et al. Characteristics, risk factors and mortality of stroke patients in kyoto, japan. BMJ open. 2013;3:e002181

5. Shigematsu K, Nakano H, Watanabe Y. The eye response test alone is sufficient to predict stroke outcome—reintroduction of japan coma scale: A cohort study. BMJ open. 2013;3:e002736

6. Shigematsu K, Nakano H, Watanabe Y, Sekimoto T, Shimizu K, Nishizawa A, et al. Speech disturbance at stroke onset is correlated with stroke early mortality. BMC neurology. 2013;13:1–7

7. Goldstein LB, Adams R, Becker K, Furberg CD, Gorelick PB, Hademenos G, et al. Primary prevention of ischemic stroke: A statement for healthcare professionals from the stroke council of the american heart association. Stroke. 2001;32:280–299

8. Goldstein LB, Adams R, Alberts MJ, Appel LJ, Brass LM, Bushnell CD, et al. Primary prevention of ischemic stroke: A guideline from the american heart association/american stroke association stroke council: Cosponsored by the atherosclerotic peripheral vascular disease interdisciplinary working group; cardiovascular nursing council; clinical cardiology council; nutrition, physical activity, and metabolism council; and the quality of care and outcomes research interdisciplinary working group: The american academy of neurology affirms the value of this guideline. Stroke. 2006;37:1583–1633

9. Rival J, Riddle JM, Stein PD. Effects of chronic smoking on platelet function. Thrombosis Research. 1987;45:75–85

10. Benowitz NL, Burbank AD. Cardiovascular toxicity of nicotine: Implications for electronic cigarette use. Trends in cardiovascular medicine. 2016;26:515–523

11. Burke A, FitzGerald GA. Oxidative stress and smoking-induced vascular injury. Progress in cardiovascular diseases. 2003;46:79–90

12. Kurth T, Kase CS, Berger K, Gaziano JM, Cook NR, Buring JE. Smoking and risk of hemorrhagic stroke in women. Stroke. 2003;34:2792–2795

13. Kurth T, Kase CS, Berger K, Schaeffner ES, Buring JE, Gaziano JM. Smoking and the risk of hemorrhagic stroke in men. Stroke. 2003;34:1151–1155

